# Waning antibody levels after vaccination with mRNA BNT162b2 and inactivated CoronaVac COVID-19 vaccines in Hong Kong blood donors

**DOI:** 10.1101/2021.12.05.21267330

**Authors:** Shirley LL Kwok, Samuel MS Cheng, Jennifer NS Leung, Kathy Leung, Cheuk-Kwong Lee, JS Malik Peiris, Joseph T Wu

## Abstract

Both inactivated vaccine (CoronaVac; Sinovac) and mRNA vaccine (Comirnaty/BNT162b2; Fosun-Pharma/BioNTech) are available in Hong Kong’s COVID-19 Vaccination Programme. We reported waning antibody levels by enzyme-linked immunosorbent assays (ELISA) and surrogate virus neutralization test (sVNT) among 850 fully vaccinated blood donors (i.e., received two doses). The BNT162b2 group’s antibody levels remain over the 50% protection threshold within six months, and the CoronaVac’s group’s median antibody levels begin to fall below the 50% protection threshold two months after vaccination.

---

The Hong Kong government launched its COVID-19 Vaccination Programme on 26 February 2021. The programme is free for all Hong Kong residents with both inactivated vaccine (CoronaVac; Sinovac) and mRNA vaccine (Comirnaty/BNT162b2; Fosun-Pharma/BioNTech) being available. As of 31 October 2021, 9,043,407 doses have been administered and over 60% (4,425,382) of the eligible population have received two doses [1].

As part of a community-based COVID-19 sero-epidemiological study, we recruited 14,169 healthy blood donors at the Hong Kong Red Cross Blood Transfusion Service from April 2020 to October 2021. Among these donors, 850 were fully vaccinated (at least 14 days after the second dose, according to the Hong Kong Centre for Health Protection) with either BNT162b2 or CoronaVac and provided their vaccination history. Of the 850 fully vaccinated blood donors, 593 (69.8%) received two doses of BNT162b2 and 257 (30.2%) received two doses of CoronaVac. The CoronaVac group was older than the BNT162b2 group (median age 48 vs 39, *p* < 0.001). The duration from receiving the second dose of vaccine to the blood donation was shown in Supplementary table 1.

All samples collected were first tested using an enzyme-linked immunosorbent assay (ELISA), which detects antibodies to the receptor-binding domain (RBD) of the severe acute respiratory syndrome coronavirus 2 (SARS-CoV-2) spike protein [2]. ELISA-positive samples were further tested using surrogate virus neutralization test (sVNT), which measures neutralising antibodies that inhibit interaction between ACE-2 human cell surface receptor and SARS-CoV-2 spike protein RBD [3]. The manufacturers’ recommended threshold for a positive sVNT test was ≥30% inhibition of signal. We arbitrarily assigned an sVNT signal inhibition percentage of 20% to ELISA-negative samples with no sVNT results, since we expect these samples to show low but non-zero result if they had been tested.

Participants were categorised by the vaccine they received, and the number of months between their vaccination and donation (from month 0 to month 6). The BNT162b2 group had a higher percentage of positive ELISA (99.2% vs. 73.2%, *p* < 0.001) and sVNT (99.0% vs. 70.4%, *p* < 0.001) results than the CoronaVac group overall. The BNT162b2 vaccinees show a generally higher level of antibodies than the CoronaVac vaccinees in both ELISA (1.94 vs. 0.86, *p* < 0.001) and sVNT (94% vs 39%, *p* < 0.001); this trend is observed from within a month after vaccination to five months after vaccination (Supplementary table 2).

Recent studies showed that a 50% plaque reduction neutralisation test (PRNT_50_) titre of 1:25.9, which is approximately 20% of the mean convalescent neutralisation level, provides 50% protection against infection of SARS-CoV-2 [4]. Since PRNT_50_ is correlated with sVNT results, we estimate a 50% protection threshold to be 45.3% inhibition in sVNT [5, 6]. Although antibody levels decline over time for both groups, the BNT162b2 group’s antibody levels remain well over the 50% protection threshold; the CoronaVac’s group’s median antibody levels begin to fall below the 50% protection threshold two months after vaccination (Supplementary table 3, Figure). We did not derive a protection threshold for ELISA since sVNT performs better in terms of test specificity and sensitivity [7].

**Figure.**
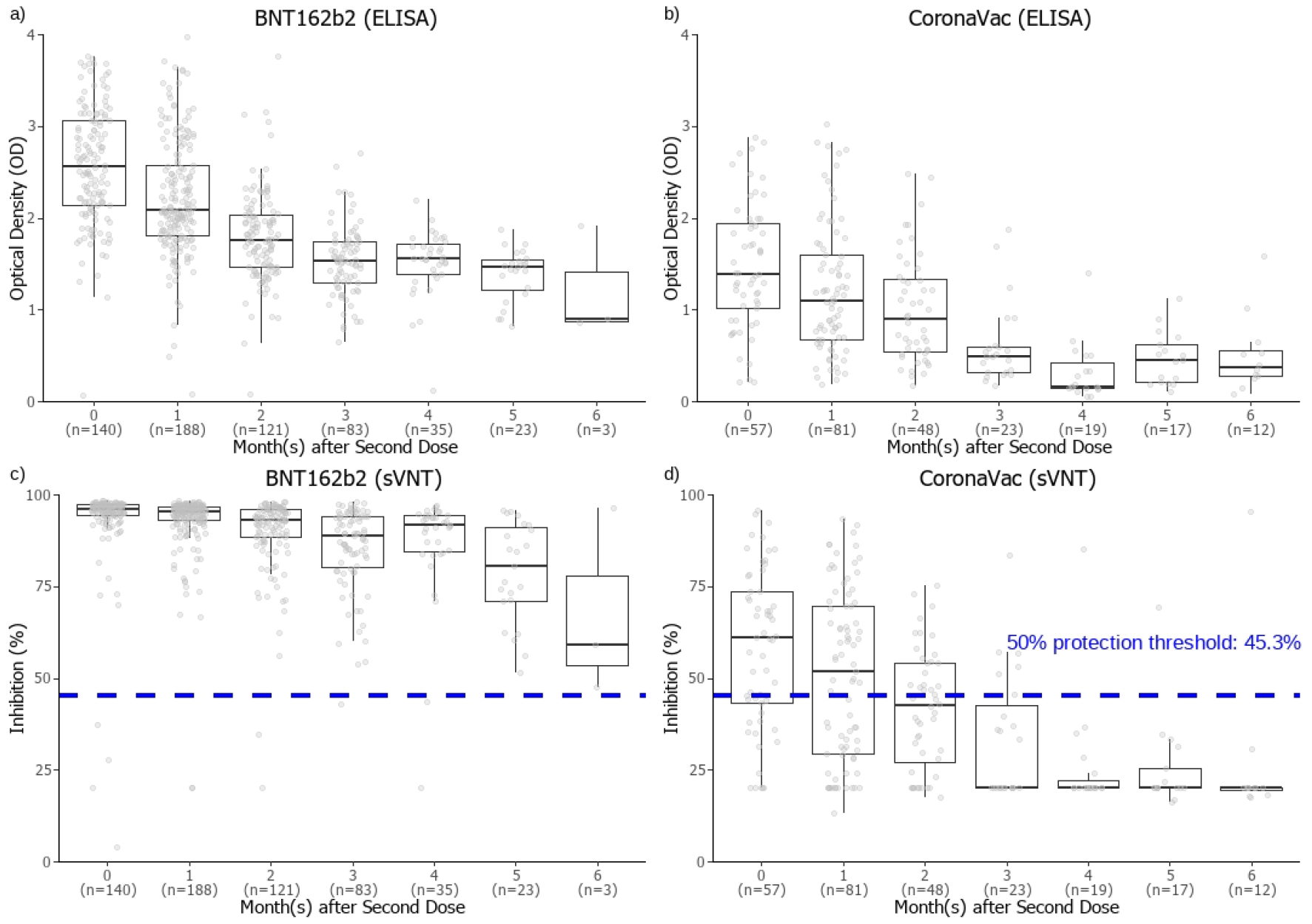
Antibody responses in Hong Kong blood donors from 14 days to 6 months after vaccination; enzyme-linked immunosorbent assay (ELISA) results for BNT162b2 vaccinees (A) and CoronaVac vaccinees (B), and surrogate virus neutralisation test (sVNT) results for BNT162b2 vaccinees (C) and CoronaVac vaccinees (D)* * Lower and upper hinges of boxplot represent first and third quartiles (interquartile range), middle hinge represents median, upper and lower whiskers extend to 1.5 times the interquartile range. Outliers are data beyond the end of the whiskers.

There have been limited studies on waning antibody levels following CoronaVac vaccine [8, 9]. Since antibody levels have been shown to be predictive of protection against SARS-CoV-2 and other variants of concern, waning antibody levels with different COVID-19 vaccines should be taken into consideration when designing vaccination programmes [3, 4, 10]. Our study did not account for other mechanisms of immune protection such as T cell response, which might also be important in assessing vaccine protection against severe infections of SARS-CoV-2 [11-13]. We also did not account for the impact of virus variants which may have reduced susceptibility to neutralizing antibody by vaccines based on the original SARS-CoV-2. Thus, our estimates are likely to be over-optimistic but provide a minimum estimate. More studies on vaccine-induced protection over time are needed to help formulate effective vaccination strategies.

## Data Availability

All data produced in the present study are available upon reasonable request to the authors.

## Acknowledgements

We would like to thank all participants from Causeway Bay, Sha Tin and Tsuen Wan Donation Centre of Hong Kong Blood Transfusion Service. We also would like to thank Ms Wing-Ching Ling and Ms Rita Lo from the Red Cross Blood Transfusion Service and Ms Miky Wong, Dr Di Liu and Ms Kitty YY Lau from the School of Public Health, University of Hong Kong for their technical support.

## Supplementary material

This supplementary material is hosted by *Eurosurveillance* as supporting information alongside the article “Waning antibody levels after vaccination with mRNA BNT162b2 and inactivated CoronaVac COVID-19 vaccines in Hong Kong blood donors” on behalf of the authors who remain responsible for the accuracy and appropriateness of the content. The same standards for ethics, copyright, attributions and permissions as for the article apply. Supplements are not edited by *Eurosurveillance* and the journal is not responsible for the maintenance of any links or email addresses provided therein.

**Supplementary table 1.** Characteristics of study cohort.

|  | Overall<br>(N = 850) |  | BNT162b2<br>(n = 593) |  | CoronaVac<br>(n = 257) |  | *p*<sup>†</sup> |
| --- | --- | --- | --- | --- | --- | --- | --- |
|  | No. of donors | % of donors | No. of donors | % of donors | No. of donors | % of donors |  |
| **Age group** |  |  |  |  |  |  | <0.001 |
| **12-19** | 16 | 1.9% | 14 | 2.4% | 2 | 0.8% |  |
| **20-29** | 137 | 16.1% | 127 | 21.4% | 10 | 3.9% |  |
| **30-39** | 199 | 23.4% | 163 | 27.5% | 36 | 14.0% |  |
| **40-49** | 243 | 28.6% | 153 | 25.8% | 90 | 35.0% |  |
| **50-59** | 195 | 22.9% | 100 | 16.9% | 95 | 37.0% |  |
| **60-69** | 60 | 7.1% | 36 | 6.1% | 24 | 9.3% |  |
| **Gender** |  |  |  |  |  |  | 0.025 |
| **Female** | 387 | 45.5% | 285 | 48.1% | 102 | 39.7% |  |
| **Male** | 463 | 54.5% | 308 | 51.9% | 155 | 60.3% |  |
| **Month(s) after vaccination** |  |  |  |  |  |  | <0.001 |
| **0** | 197 | 23.2% | 140 | 23.6% | 57 | 22.2% |  |
| **1** | 269 | 31.6% | 188 | 31.7% | 81 | 31.5% |  |
| **2** | 169 | 19.9% | 121 | 20.4% | 48 | 18.7% |  |
| **3** | 106 | 12.5% | 83 | 14.0% | 23 | 8.9% |  |
| **4** | 54 | 6.4% | 35 | 5.9% | 19 | 7.4% |  |
| **5** | 40 | 4.7% | 23 | 3.9% | 17 | 6.6% |  |
| **6** | 15 | 1.8% | 3 | 0.5% | 12 | 4.7% |  |

† Tested using Fisher's exact test.

**Supplementary table 2.** ELISA and sVNT results from 14 days to 6 months after vaccination, comparison between BNT162b2 and CoronaVac*.

|  | Month 0 |  |  |  |  | Month 1 |  |  |  |  |
| --- | --- | --- | --- | --- | --- | --- | --- | --- | --- | --- |
|  | BNT162b2<br>(n = 140) |  | CoronaVac<br>(n = 57) |  | *p*<sup>†</sup> | BNT162b2<br>(n = 188) |  | CoronaVac<br>(n = 81) |  | *p*<sup>†</sup> |
|  | % | 95% CI | % | 95% CI |  | % | 95% CI | % | 95% CI |  |
| **ELISA** |  |  |  |  |  |  |  |  |  |  |
| % of Positives<br>(≥0.5) | 99.3% | 96.1-100 | 91.2% | 80.7-97.1 | 0.008 | 98.9% | 96.2-99.9 | 85.2% | 75.6-92.1 | <0.001 |
| **sVNT** |  |  |  |  |  |  |  |  |  |  |
| % of Positives<br>(≥30%) | 98.6% | 94.9-99.8 | 91.2% | 80.7-97.1 | 0.022 | 98.9% | 96.2-99.9 | 84.0% | 74.1-91.2 | <0.001 |
| % above 50%<br>protection<br>threshold<br>(≥45.3%) | 97.1% | 92.8-99.2 | 66.7% | 52.9-78.6 | <0.001 | 98.9% | 96.2-99.9 | 56.8% | 45.3-67.8 | <0.001 |

|  | Month 2 |  |  |  |  | Month 3 |  |  |  |  |
| --- | --- | --- | --- | --- | --- | --- | --- | --- | --- | --- |
|  | BNT162b2<br>(n = 121) |  | CoronaVac<br>(n = 48) |  | *p*<sup>†</sup> | BNT162b2<br>(n = 83) |  | CoronaVac<br>(n = 23) |  | *p*<sup>†</sup> |
|  | % | 95% CI | % | 95% CI |  | % | 95% CI | % | 95% CI |  |
| **ELISA** |  |  |  |  |  |  |  |  |  |  |
| % of Positives<br>(≥0.5) | 99.2% | 95.5-100 | 79.2% | 65.0-89.5 | <0.001 | 100% | 95.7-100 | 47.8% | 26.8-69.4 | <0.001 |
| **sVNT** |  |  |  |  |  |  |  |  |  |  |
| % of Positives<br>(≥30%) | 99.2% | 95.5-100 | 77.1% | 62.7-88.0 | <0.001 | 100% | 95.7-100 | 47.8% | 26.8-69.4 | <0.001 |
| % above 50%<br>protection<br>threshold<br>(≥45.3%) | 98.3% | 94.2-99.8 | 43.8% | 29.5-58.8 | <0.001 | 98.8% | 93.5-100 | 26.1% | 10.2-48.4 | <0.001 |

|  | Month 4 |  |  |  |  | Month 5 |  |  |  |  |
| --- | --- | --- | --- | --- | --- | --- | --- | --- | --- | --- |
|  | BNT162b2<br>(n = 35) |  | CoronaVac<br>(n = 19) |  | *p*<sup>†</sup> | BNT162b2<br>(n = 23) |  | CoronaVac<br>(n = 17) |  | *p*<sup>†</sup> |
|  | % | 95% CI | % | 95% CI |  | % | 95% CI | % | 95% CI |  |
| **ELISA** |  |  |  |  |  |  |  |  |  |  |
| % of Positives<br>(≥0.5) | 97.1% | 85.1-99.9 | 26.3% | 9.1-51.2 | <0.001 | 100% | 85.2-100 | 47.1% | 23.0-72.2 | <0.001 |
| **sVNT** |  |  |  |  |  |  |  |  |  |  |
| % of Positives<br>(≥30%) | 97.1% | 85.1-99.9 | 26.3% | 9.1-51.2 | <0.001 | 100% | 85.2-100 | 35.3% | 14.2-61.7 | <0.001 |
| % above 50%<br>protection<br>threshold<br>(≥45.3%) | 94.3% | 80.8-99.3 | 5.3% | 0.1-26.0 | <0.001 | 100% | 85.2-100 | 5.9% | 0.1-28.7 | <0.001 |

|  | Month 6 |  |  |  |  | Overall |  |  |  |  |
| --- | --- | --- | --- | --- | --- | --- | --- | --- | --- | --- |
|  | BNT162b2<br>(n = 3) |  | CoronaVac<br>(n = 12) |  | *p*<sup>†</sup> | BNT162b2<br>(n = 593) |  | CoronaVac<br>(n = 257) |  | *p*<sup>†</sup> |
|  | % | 95% CI | % | 95% CI |  | % | 95% CI | % | 95% CI |  |
| **ELISA** |  |  |  |  |  |  |  |  |  |  |
| % of Positives<br>(≥0.5) | 100% | 29.2-100 | 41.7% | 15.2-72.3 | 0.2 | 99.2% | 98.0-99.7 | 73.2% | 67.3-78.5 | <0.001 |
| **sVNT** |  |  |  |  |  |  |  |  |  |  |
| % of Positives<br>(≥30%) | 100% | 29.2-100 | 16.7% | 2.1-48.4 | 0.022 | 99.0% | 97.8-99.6 | 70.4% | 64.4-75.9 | <0.001 |
| % above 50%<br>protection<br>threshold<br>(≥45.3%) | 100% | 29.2-100 | 8.3% | 0.2-38.5 | 0.009 | 98.1% | 96.7-99.1 | 44.4% | 38.2-50.7 | <0.001 |

\* 95% confidence interval calculated from binomial distribution using the Clopper-Pearson method.
† Tested using Fisher's exact test.

**Supplementary table 3a.** ELISA and sVNT results from 14 days to 6 months after vaccination with BNT162b2*.

|  | BNT162b2 |  |  |  |  |  |  |  |  |  |  |  |  |  |  |
| --- | --- | --- | --- | --- | --- | --- | --- | --- | --- | --- | --- | --- | --- | --- | --- |
|  | Month 0<br>(n = 140) |  | Month 1<br>(n = 188) |  | Month 2<br>(n = 121) |  | Month 3<br>(n = 83) |  | Month 4<br>(n = 35) |  | Month 5<br>(n = 23) |  | Month 6<br>(n = 3) |  | *p*† |
|  | % | 95% CI | % | 95% CI | % | 95% CI | % | 95% CI | % | 95% CI | % | 95% CI | % | 95% CI |  |
| **ELISA** |  |  |  |  |  |  |  |  |  |  |  |  |  |  |  |
| % of Positives (≥0.5) | 99.3% | 96.1-100 | 98.9% | 96.2-99.9 | 99.2% | 95.5-100 | 100% | 95.7-100 | 97.1% | 85.1-99.9 | 100% | 85.2-100 | 100% | 29.2-100 | 0.7 |
| **sVNT** |  |  |  |  |  |  |  |  |  |  |  |  |  |  |  |
| % of Positives (≥30%) | 98.6% | 94.9-99.8 | 98.9% | 96.2-99.9 | 99.2% | 95.5-100 | 100% | 95.7-100 | 97.1% | 85.1-99.9 | 100% | 85.2-100 | 100% | 29.2-100 | 0.7 |
| % above 50% protection threshold (≥45.3%) | 97.1% | 92.8-99.2 | 98.9% | 96.2-99.9 | 98.3% | 94.2-99.8 | 98.8% | 93.5-100 | 94.3% | 80.8-99.3 | 100% | 85.2-100 | 100% | 29.2-100 | 0.2 |

\* 95% confidence interval calculated from binomial distribution using the Clopper-Pearson method.
† Tested using Fisher's exact test.

**Supplementary table 3b.** ELISA and sVNT results from 14 days to 6 months after vaccination with CoronaVac*.

|  | CoronaVac |  |  |  |  |  |  |  |  |  |  |  |  |  |  |
| --- | --- | --- | --- | --- | --- | --- | --- | --- | --- | --- | --- | --- | --- | --- | --- |
|  | Month 0<br>(n = 57) |  | Month 1<br>(n = 81) |  | Month 2<br>(n = 48) |  | Month 3<br>(n = 23) |  | Month 4<br>(n = 19) |  | Month 5<br>(n = 17) |  | Month 6<br>(n = 12) |  | *p*† |
|  | % | 95% CI | % | 95% CI | % | 95% CI | % | 95% CI | % | 95% CI | % | 95% CI | % | 95% CI |  |
| **ELISA** |  |  |  |  |  |  |  |  |  |  |  |  |  |  |  |
| % of Positives (≥0.5) | 91.2% | 80.7-97.1 | 85.2% | 75.6-92.1 | 79.2% | 65.0-89.5 | 47.8% | 26.8-69.4 | 26.3% | 9.1-51.2 | 47.1% | 23.0-72.2 | 41.7% | 15.2-72.3 | <0.001 |
| **sVNT** |  |  |  |  |  |  |  |  |  |  |  |  |  |  |  |
| % of Positives (≥30%) | 91.2% | 80.7-97.1 | 84.0% | 74.1-91.2 | 77.1% | 62.7-88.0 | 47.8% | 26.8-69.4 | 26.3% | 9.1-51.2 | 35.3% | 14.2-61.7 | 16.7% | 2.1-48.4 | <0.001 |
| % above 50% protection threshold (≥45.3%) | 66.7% | 52.9-78.6 | 56.8% | 45.3-67.8 | 43.8% | 29.5-58.8 | 26.1% | 10.2-48.4 | 5.3% | 0.1-26.0 | 5.9% | 0.1-28.7 | 8.3% | 0.2-38.5 | 0.002 |

\* 95% confidence interval calculated from binomial distribution using the Clopper-Pearson method.
† Tested using Fisher's exact test.

